# Impact of small sample sizes on the internal quality control: statistical uncertainties in the determination of root mean square deviations with respect to the sample mean (RMSD) or to a target value (RMSTD)

**DOI:** 10.1101/2020.12.10.20247148

**Authors:** Christian Beier

## Abstract

Insufficient statistics due to small considered sample sizes can cause distinct problems in internal quality control (IQC) approaches. This issue concerns most of the currently applied IQC concepts either directly (if a root-mean-square-deviation metric is evaluated) or indirectly (if the IQC concept facilitates a standard deviation that was self-evaluated based on a very limited number (n≤30) of control measures). In clinical chemistry a famous example for the latter case is the common implementation of the Westgard Sigma Rules approach.

This study quantifies the statistical uncertainties in the determination of root mean square (total) deviations related to the sample mean (RMSD) or to a target value (RMSTD). It is clearly shown that RMS(T)D values based on small data sets with n<50 samples are accompanied by a significant statistical uncertainty that needs to be considered in adequate IQC limit definitions. Two mathematical models are derived to reliably estimate an optimal adaptation function to adjust IQC limits to short charts of control measures.

This article provides the theoretical background for the novel IQC method “Statistical Monitoring by Adaptive RMSTD Tests” (SMART) intended to monitor limited available numbers of recent control measures (usually n<20). The study also addresses a general problem in specificity of an IQC resulting from too small sample sizes during the evaluation period of the applied in-control standard deviation.

## 1 Introduction

Quality assurance, and particularly the internal quality control (IQC), are essential requirements to monitor the reliability of processes in industry and health care. An IQC of quantitative methods usually utilizes control sample(s), which are continuously captured. Aside from evaluations of each Single Measurement of a Control sample (SMC values), a statistical retrospective analysis (RA) of recent SMC data allows to early identify changes in accuracy and precision of the measuring system. Efficient and widely used statistical metrics are the root mean square deviation RMSD (aka standard deviation) and the root mean square total deviation RMSTD. The latter refers to a given target value of the control sample instead of the current average.

Based on a chart of consecutive SMC values, the current standard deviation s_n_ and the unsigned bias (mean inaccuracy) δ_n_ can be revealed by

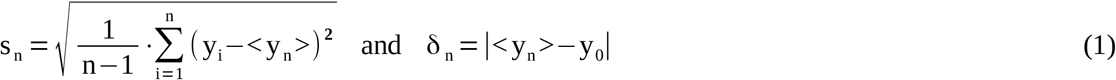

(y_i_: measured SMC, y_0_: target value, <y_n_>: sample mean, n: sample size of an SMC chart). Particularly in the case of low-frequent sampling, the parameters s_n_ and δ_n_ are not strictly separable because:

i. The measurement inaccuracy (bias) can vary even within a single chart due to unpreventable or tolerated changes in environmental or operating conditions. These shifts contribute to the amount of the standard deviation.
ii. The average values of arbitrary SMC charts of length n scatter themselves with a dispersion of 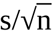. Thus, the amount of the standard deviation affects the obtained average value of a particular chart of limited size - leading to an arbitrary distortion of the real bias of this chart. It is therefore suitable to solely focus on the entire analytical measuring uncertainty (aka RMSTD)

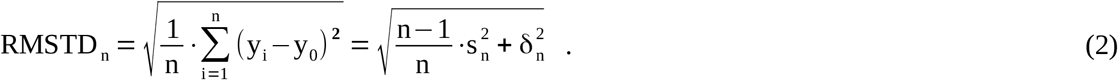

The RA has to handle a trade-off between (i) a sufficient number of SMC samples to reliably verify a statistical in-control condition and (ii) a special focus on most recent data to detect changes in measuring precision as fast as possible. Particularly in clinical chemistry, the RA has to deal with generally low-frequent SMC measures (1-2 SMC per day and control sample) as well as limited reasonable collection periods due to short reagent lifetimes or other frequent system interventions. Thus, the considered SMC data set is often very small on a statistical point of view. Some prescribed RA approaches in national guidelines demand only n=15 retrospective values [1]. This small number has consequences for the applied maximum permissible limits of statistical metrics, which are often defined for “sufficiently long” charts of SMC data (i.e., with little or no consideration of remaining statistical uncertainties in determination). The minimum amount of collected SMC data to reliably apply such limits is generally significantly underestimated. To prevent false out-of-control alerts, general limits have to be adapted to account for the increased statistical uncertainty at very limited sample sizes. We thus need to know, how an adequate RMSTD (or RMSD) limit level L(n) must be increased when it is applied to smaller and smaller sample sizes. The functional dependency of L(n) on n is the main topic of this article. In particular, the necessary adaptation function a(n) defined by L(n)=a(n)·L(∞) is investigated, where a(n) is equivalent to the relative limit L_rel_(n)=L(n)/L(∞) and L_rel_(n→∞)=1. The adaptation function of the RMSTD metric serves as an integral part of the novel and powerful IQC method “Statistical Monitoring by Adaptive RMSTD Tests (SMART)” [2]. This method efficiently covers all basic aspects of an IQC, and it is particularly suitable for low-frequent sampling. Alternatively, the provided results may facilitate a new type of confirmed RMSD-based short-term control rules, applicable in other multirule IQC concepts.

All limits defined in this article are unsigned values specifying a range around the target value of the control sample y_0_±L_given_.

The “Westgard Sigma Rules” [3-5] is currently the most prominent IQC approach in clinical chemistry. It is a multirule concept of consecutively applied simple tests. The entry rule is a range test with a cut-off limit of three times the expected standard deviation around the expected mean 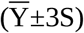. Both parameters S and 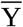 need to be determined during a pre-analytical evaluation period, if there are no explicitly prescribed values. Evaluation periods are commonly done using a very small considered sample size (n about 20). The resulting distinct statistical uncertainty implies a high risk of significantly underestimated amounts of S and 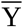. This problem and the impact to the Westgard Sigma Rules will be quantified and discussed in Chapter 3.4 of this article.

## 2 Methods

### 2.1 The determination of the “true” RMSTD

The abbreviation Δ will be used for simplicity to denote the (almost) exactly determined “true” RMSTD value, which can be revealed by a sufficiently large SMC data set (n>>100). The word “true” emphasizes that there is no remaining statistical uncertainty in the determination of the RMSTD, standard deviation, or mean bias.

The evaluated Δ value of a control sample also defines the smallest permissible limit of the RMSTD (based on and intended for large SMC data). Thus, the Δ value should nevertheless include all kinds of measuring uncertainties that are unpreventable or tolerable during the in-control operation state of the measuring system. Ideally, the determination of Δ is required for each particular combination of a control material and reagent lot. However, it is a common practice to accept a certain level of generalization. In special cases one prescribed general limit value might be sufficient for every use of a measuring system. The determination of Δ utilizes Eq. (2) and ideally requires extensive statistics over several long SMC charts of the same control sample

- with different devices,
- under different permissible operating and environmental conditions,
- including several recalibrations, and
- using different lots (if the target value is not lot-specific).

(The entire procedure may include separate statistics of different control samples with analyte concentrations predominantly near the medical decision limit.)

The SMC survey is best realized by sharing of peer-group data. The in-control state is defined by all considered data sets. Thus, a mechanism should be applied to identify and entirely reject suspect data sets that distinctly diverge from the group averages (e.g., an ANOVA test).

The collected overall data set of SMC values still includes outliers. To ensure a robust statistics, distinct outliers should be identified and rejected. This can approximately be done under the assumption that acceptable measures (defining the in-control state) are normal-distributed around the mean, whereas outliers are independently distributed. Thus, the elimination of outliers utilizes a recurrently updated fit of the histogram over all currently accepted SMC values by a Gaussian function. Most diverging SMC values (with respect to the mean) are sorted out as long as the quality of the Gaussian fit of the remaining data set increases.

The deviation and bias of the final Gaussian fit function directly provide the essential parameters S (overall standard deviation), Y_mean_ (overall mean value), and δ_mean_ (mean bias). Hence, the true RMSTD is revealed from S and δ_mean_ based on the general relation

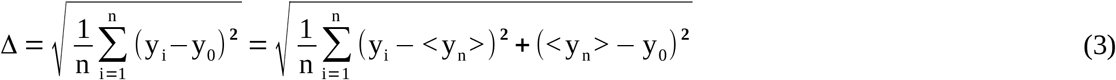

(y_i_: sample value; y_0_: target value; <y_n_>: sample mean; n: entire/large sample size), which can be transformed to

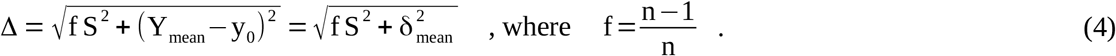

Some equations in Chapter 2.2 will utilize the following conversion rule to separate S:

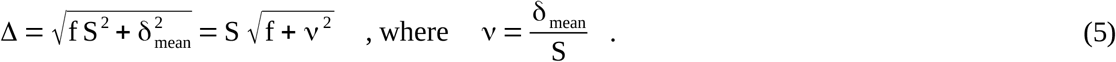

In this way, the standalone parameter S can finally be eliminated in relative RMSTD metrics. A discussion concerning the range of realistic values of ν is given in Chapter 3.1. For sufficiently large n, the factor f can be substituted by f=1; thus, Δ becomes 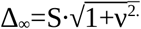.

### 2.2 The maximum error propagation of statistical uncertainties

An applied IQC limit L(n) for single-chart RMSTD values must be equal or higher than the RMSTD of any possible in-control data set of n considered SMC values. The maximally expected RMSTD value of a limited data set can be approximated by the upper limit of the n-dependent confidence interval (CI_Δ_^up^) of the RMSTD metric (at an adequate confidence level of 0.95 or higher). Here, the upper CI limit is the sum of the true value Δ and the expected statistical uncertainty u(Δ) at a given sample size n

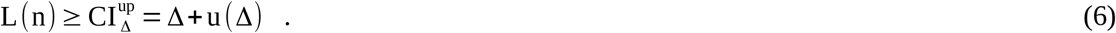

To apply reasonable limits, it is beneficial to estimate the curvature of CI_Δ_^up^ at small sample sizes. This curvature will finally provide inference to the necessary adaptation of general IQC limits for the RMSTD metric with regard to an arbitrary (limited) sample size. In cases of very large n, the statistical uncertainty u(Δ) can be neglected, leading to L(n→∞)≥Δ_∞_. The basic Eq. (4) of Δ contains two empiric variables S^2^ and Y_mean_, having an error due to their statistical uncertainties. The variance S^2^ is preferred instead of the standard deviation, because an entirely unbiased estimator with regard to different sample sizes only exists for S^2^.

To estimate u(Δ) by u(S^2^) and u(Y_mean_) the maximum error propagation (first-order Taylor series expansion) is applied, utilizing the absolute amounts of the partial derivations *∂*(Δ)/*∂*(S^2^) and *∂*(Δ)/*∂*(Y_mean_)

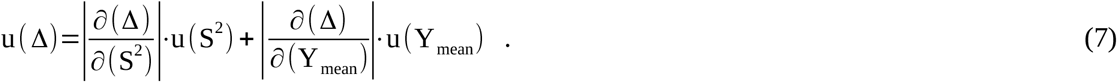

According to Eqs. (4) and (5), the required partial derivations lead to

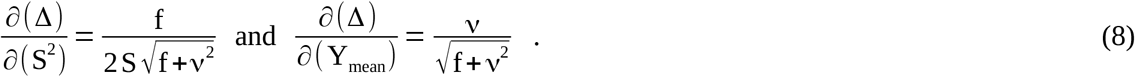

Both derivations are always positive.

The sample mean of a normal-distributed variable is itself normal-distributed with a dispersion, equal to the overall standard deviation S divided by the square root of n. Hence, the mean values <y_n_> of SMC charts of length n can be standardized according to

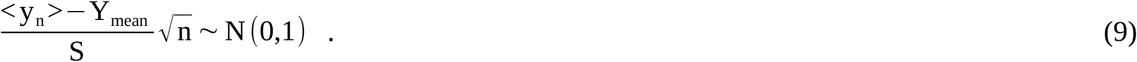

In literature, CI limits are generally given for the common case that the true parameter value of the total population (e.g., Y_mean_) is unknown. Thus, a CI is defined around the usually known value <y_n_> of a small data set, where the true value Y_mean_ is expected to be covered by the CI (at a certain confidence level). Alternatively, an equivalent (i.e., inverted) CI can also be applied to the known global variable Y_mean_, where the unknown sample mean <y_n_> is expected to lie within the inverted interval around Y_mean_ accordingly. In case of the mean, the CI definition is symmetric, and the normal and inverted definitions are identical. Though, the asymmetric CI around the global variance S^2^ (see below) differs from the inverted one around the variance s_n_^2.^ of a sample subset. Thus, following CI definitions may differ from those typically mentioned in textbooks.

In brief, the CI and uncertainty of Y_mean_ are

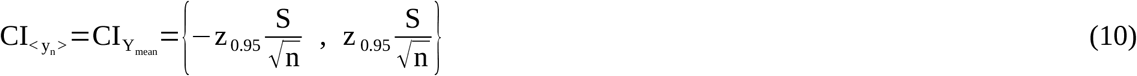

(z _0.95_ : quantile of the normal distribution with a significance level of 5%) and

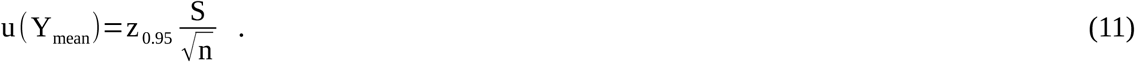

The choice of an optimal value of z_0.95_ is discussed in Chapter 3.1. The applied overall standard deviation S (revealed as described in Chapter 2.1) is a maximum estimator delimiting the general in-control state. Consequently, the expected real standard deviation of an individual SMC chart is most probably smaller because the device and measuring conditions are almost unchanged. One might argue that the student distribution t_n-1_ (instead of z) is favorable in this case, which would result in much higher amplitudes at very small n. Though, the uncertainty of S is already explicitly considered in a separate term (see Eq. (7)). The substitution of z by t_n-1_ would thus clearly overestimate u(Y_mean_). It would further lead to extremely artificial results of Eq. (29) for n<3.

The uncertainty of the overall variance S^2^ utilize the chi-squared distribution χ^2^

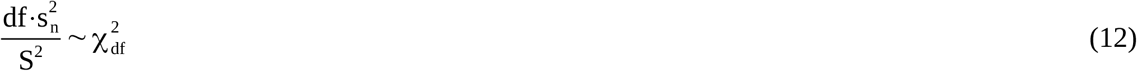

(df: degrees of freedom; s_n_^2^: variance of a sample subset of size n; χ^2^: chi-squared distribution). Thus, (at a probability of 1-α) the expected variance of an arbitrary n-sized sample series lies within the CI around the “true” variance S^2^ defined by

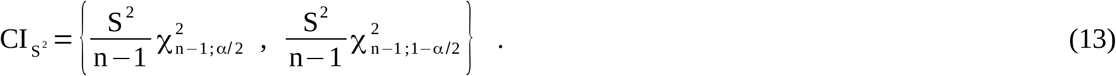

The uncertainty of the overall variance is consequently

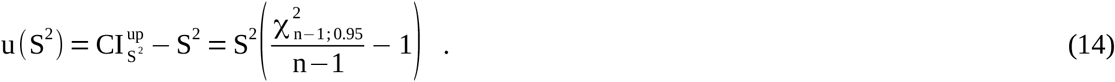

Only the upper limit of S^2^ is relevant for our purpose; thus, a one-sided confidence level (1-α/2=0.95) is used.

A particular difficulty of the u(S^2^) term of the CI_Δ_^up^ approach is the question, if the expected sample mean can be considered as known (i.e., equal to the overall Y_mean_). The actual true mean value (or true bias) of a realistic SMC chart depends on the specific device, lot, and other measuring conditions. The variation of the true means of SMC charts at different measuring conditions could be significant. It is assumed that Y_mean_ (based on a peer-group survey) can not be taken as an ideal estimate for the expected mean of a particular chart. Further, u(S^2^) should specify the error of the pure dispersion of a single chart, where the uncertainty of the contribution |<y_n_>-y_0_|^2^ is considered separately (see Eqs. (3) and (7)). Thus, Eq. (13) is intended for an unknown expectation value. This means that the unknown sample mean <y_n_> is taken as the most suitable reference for the expectation value so s_n_^2^=1**/**(n-1)*Σ_i→n_ (y_i_ -<y_n_ >)^2^. Consequently, the number of the degrees of freedom is n reduced by one (df=n-1 in Eq. (12)). This decision might be controversial. One may argue that Y_mean_ relates to S; thus, it is always a proper estimate for the expectation value of any subset mean. This would lead to df=n instead. The latter case requires that the ratio χ^2^(n-1,0.95)/(n-1) would have to be replaced by χ^2^(n,0.95)/n in Eq. (13) and in all subsequent equations. The topic is more complex, which is also reflected by the fact that the uncertainty u(S^2^) is always higher in the case of df=n-1 than in the alternative case with df=n. Here, this issue leads to an intrinsic “inexactness” of the given mathematical model, which has been visualized by function corridors in several figures. The preferred variant given in Eq. (13) indicates the upper limit of the corridor, where the lower limit is defined by the alternative case with χ^2^(n,0.95)/n. However, it should again be mentioned that the overall S is a maximum estimate of the expected standard deviation of a single chart (due to the multi-chart origin of S). Nevertheless, at a CI of 95% and no considered bias (ν=0), the preferred variant shows best agreement with the MDCI function (see Chapter 3.2).

Combining Eqs. (7), (8), (11), and (14), the entire uncertainty of Δ can now be expressed as

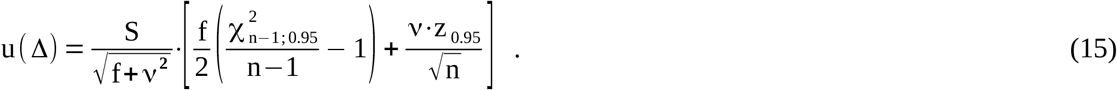

According to Eq. (6), the requested upper confidence limit of the total measuring uncertainty under poor statistical conditions is finally given by

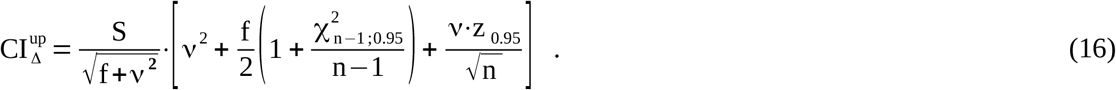

(n: number of considered SMC values, ν: ratio between δ_mean_ and S, f=(n-1)/n, χ^2^ and z: quantiles of the chi-squared and normal distributions).

The given mathematical approach is intended to estimate the statistical uncertainty of an RMSTD of small series. However, the presented mathematical model becomes unreliable for any n below about 6, due to (i) the approximative concept of the degrees of freedom (esp. the correction factor f) and (ii) the fact that the underlying Taylor-series development is limited to the first degree, which is not intended for large uncertainties. Thus, a more fundamental (but also more specific) approach is presented in the next chapter to further investigate the amount of the statistical uncertainty at very small sample sizes.

### 2.3 The Multidimensional Confidence Interval

The following derivation neglects the mean bias to the target value of the control material. Thus, it focuses on the statistical uncertainty of the RMSD (aka empirical standard deviation) in cases of small sample sizes n. As given in Eq. (12), the squared relation between a particular empirical standard deviation s_n_ of a sample and the true standard deviation S_0_ follows a chi-squared distribution. Here, a straight-forward derivation of the statistical uncertainty of RMSD values will be presented, which is comparable but not identical to the derivation of the chi-square function (which is outlined in relevant textbooks).

This fundamental theory extends the CI of single measures to a collective CI of a series of n measures, while keeping the overall confidence level constant (at, e.g., 95%). It will be denoted as the Multi-Dimensional Confidence Interval (MDCI). Hereafter, it is assumed that all (in-control) measures of the control sample are realizations of a normal-distributed totality, and each value had been standardized in advance. The MDCI theory is therefore dedicated to realizations of the standard Gaussian distribution with RMSD(n→∞)=1. Further, all SMC measures are treated to be uncorrelated to simplify the theory.

A standardization requires the knowledge of the true mean value and the true standard deviation of the particular SMC chart, which are unknown. Applying the approach described in Chapter 2.1, the overall parameters Y_mean_ and S can be revealed instead. By taking Y_mean_ as the mean value for standardization, the MDCI theory neglects the mean amount of the bias. Under the assumption that the actual bias of a limited SMC chart is variable and normal-distributed around Y_mean_, this variable fraction would be almost indistinguishable from pure statistical imprecision on a global view (see Chapter 2.1). The estimator S also considers this fraction; thus, S_0_ is overestimated by S. Fortunately, we are interested in maximum estimators to define upper limits, consequently S can be treated as an acceptable substitution for S_0_. However, it has to be mentioned that the additional contribution of the variation of the bias to S_0_, which is low-frequent, may lead to a noticeable short-term correlation between consecutive SMC measures.

The two-sided confidence interval z_95_ for a single SMC measure y is given by the integration limit that covers 95% of the area under the normalized Gaussian probability density (GPD) function

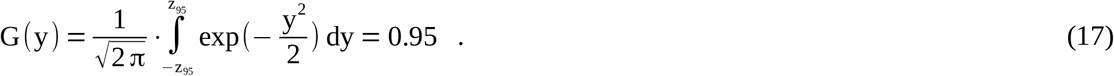

To define a confidence interval for a complete series of n measures, G must be extended to n dimensions. This is easily understandable by the general fact that the entire probability of a sequence of measures equals to the product of the unique probabilities of each measure. Here, scalar probabilities are just extended to probability distribution functions. Due to

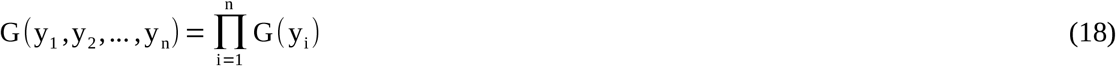

it does not matter if the measures occur in concert (multivariate) or sequentially (sample chart). Thus, the multidimensional representation of G equals to the known multivariate Gaussian distribution

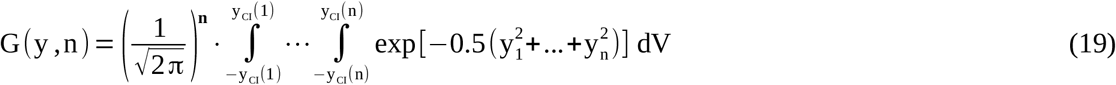

(y_CI_(n): one-dimensional confidence limits; dV: finite Cartesian volume element), where each dimension represents one measure of the SMC sample series. G(y,n) is fully rotationally symmetric around zero, thus best represented in polar co-ordinates by the radius and n-1 orientation angles. Here, we are only interested in delimiting equipotential spherical hypersurfaces of G(y,n) without further angular dependency. Hence, after pre-integration over the entire angular ranges, G(y,n) can be expressed as a normalized radial function

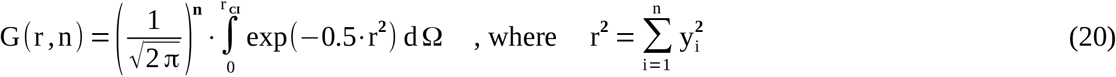

(r _CI_ : radial n-dimensional confidence limit; dΩ: finite spherical volume element in n dimensions). If G(r,n) is integrated over the radius r (origin at zero) of the Gaussian distribution in n dimensions, the size of the finite spherical volume element dΩ increases according to the area of the hypersurface of an n-dimensional sphere [6-8]. Thus, dΩ is the entire surface area of an n-dimensional sphere of radius r multiplied by a finite thickness dr

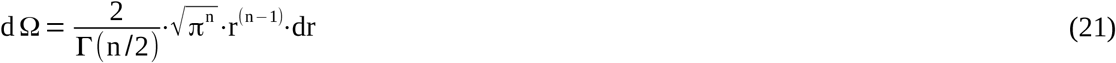

(Γ(n/2): value of the gamma function for n over two).

The values of the gamma function are usually tabulated for small n (see also Table S1 on last page). However, this approach may require values beyond the size of common gamma tables. The following set of equations can be used to reveal any requested value of gamma

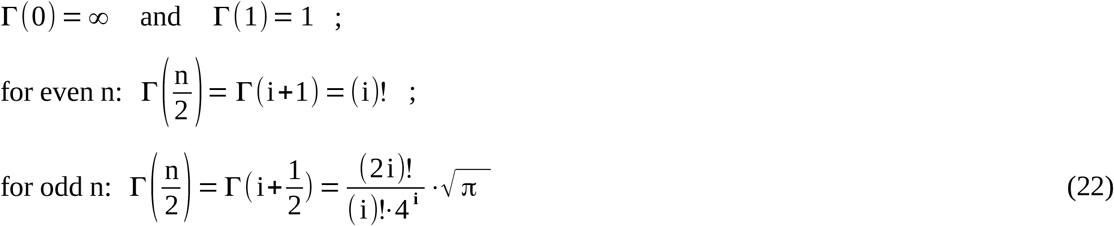

(i: integer number; “(i)!”: denotes the factorial of i).

The combination of Eqs. (20) and (21) leads to the final expression of the volume integral over the radial Gaussian probability density function in n dimensions

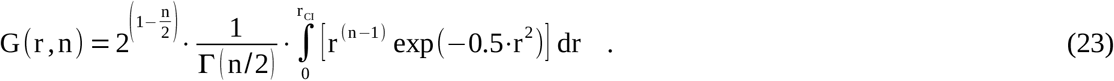

G(r,n) is normalized, which has been approved by both numeric and algebraic integration. To algebraically validate that G equals 1 over the entire range, the integral must be made consistent with a known indefinite integral. The best candidate is the general definition of the gamma function

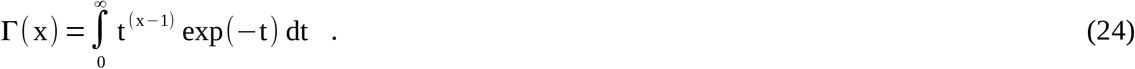

A suitable conformity to the integral term of Eq. (23) can be achieved by substitution with t=0.5·r^2^ in Eq. (23). Under consideration of following relations

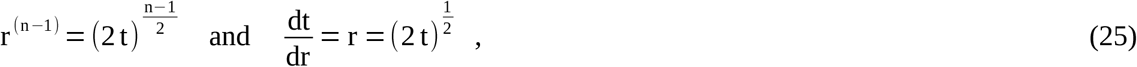

the normalization factor C of the integral term of Eq. (23) can be revealed by

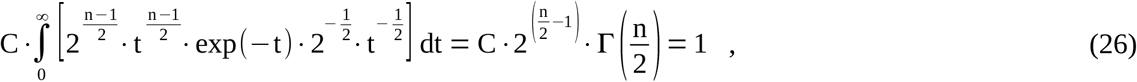

where Eq. (24) has been applied with x=n/2. After rearrangement, C in Eq. (26) is in full agreement with the normalization factor of Eq. (23).

To obtain the common (two-sided) 95% confidence limit z_95_(n) of a series of n measures, the normalized radial density function (Eq. (23)) has to be integrated from 0 to z_95_(n)

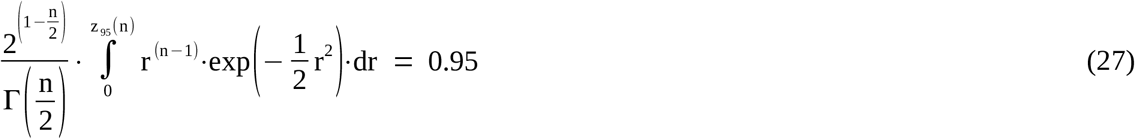

(n: sample size or dimension; r: radius of the spherical partitioning hypersurface; Γ: Gamma function). Thus, the radial integration limit r_CI_=z_95_(n) covers 95% of the hypervolume spanned by the multidimensional GPD distribution.

Obviously, z_95_ is 1.96 at n=1. For higher n, the limit z_95_(n) increases according to Eq. (27) and slowly converges towards the square root function of n. According to Eq. (20), the limit z_95_(n) represents the Euclidean distance r_CI_ of a series of single confidence limits ensuring a constant overall significance level of 5%. The limit can therefore be utilized to obtain the maximum acceptable RMSD (with 95% confidence) of a sequence of n SMC measures. To obtain such a function of adequate upper RMSD limits L°(n) of n standardized values, z_95_(n) can directly be implemented into Eq. (2) (considering y_0_=0) leading to

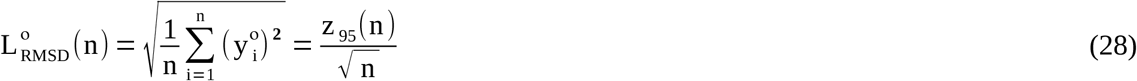

(° denotes the RMSD limit of an N(0,1)-distributed sample; y_i_°: standardized SMC values). L°(n) goes from 1.96 (n=1) to 1 (for n→∞) as drawn in Fig. 1. To apply L°(n) to non-standardized y_i_ values, L°(n) has to be back-transformed using the overall standard deviation S and Y_mean_, thus L_RMSD_(n)=S·L°(n)+Y_mean_.

**Figure 1:**
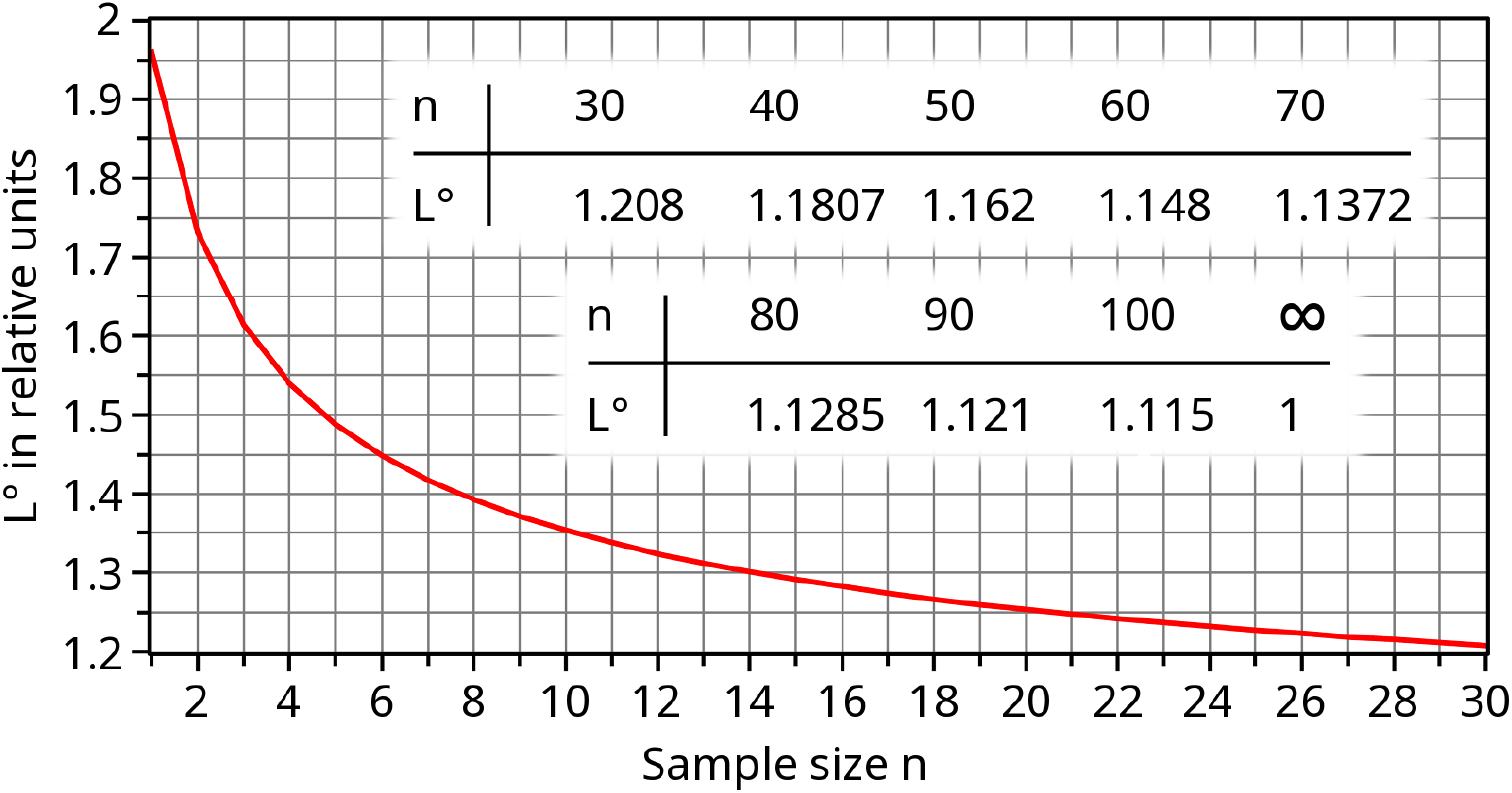
The shape of L° (Eq. (28)) of the MDCI theory is provided graphically (n≤30) and tabularly (n≥30).

## 3 Results and conclusions

### 3.1 The uncertainty of the RMSTD at small sample sizes

To provide a general curvature of the increase of the statistical uncertainty at smaller sample sizes, the CI_Δ_^up^(n) (given in Eq. (16)) is normalized by the true RMSTD value for n→∞, which is 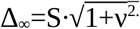 as defined at the very end of Chapter 2.1. It leads to the relative adaptation function for RMSTD limits

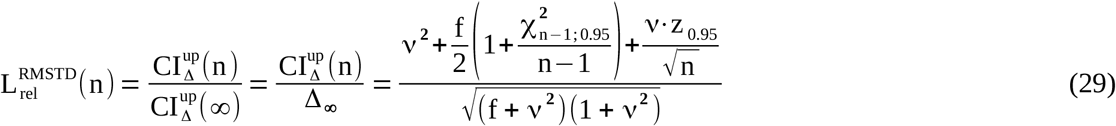

(n: sample size; f=(n-1)/n; ν=δ_mean_/S; χ^2^ and z: 95%-quantiles of the chi-squared and normal distributions). The relative CI_Δ_^up^ function L_rel_^RMSTD^ depends on the ratio ν between bias and standard deviation but not on the absolute amount of S.

Although an important goal of measuring techniques should be the minimization of a bias (e.g., by advanced non-linear calibration), the actual biases of current analytic systems in clinical chemistry are often relevant. It is important to mention that the ratio ν of the overall parameters δ_mean_ and S is not directly comparable to the ratio of the maximum permissible values δ_max_ and s_max_ intended for single SMC charts. The IQC limits δ_max_ and s_max_ issue from a (deprecated) separate evaluation of inaccuracy and imprecision, where δ_max_ restricts the permissible variability of the bias of SMC charts. The individual biases (quantified by the particular average value) of single SMC charts vary according to device-specific, operational, and environmental conditions with a scatter parameter S_δ_. The procedure to obtain δ_mean_ and S, presented in Chapter 2.1, includes SMC data from a large amount of SMC charts. In this overall statistics, the variation of the biases of individual charts is thus part of the overall standard deviation term S. The implicitly included variation of biases can be treated as normal-distributed. Pure short-term dispersion (true statistical imprecision) and the variation of biases are further sufficiently independent; thus, the overall variance is the sum of both variances S^2^=S^2^_imp_ +S^2^_δ_. The remaining overall bias δ is just the difference between the overall mean value and the target value. If the target value is a nominal value, δ_mean_ is expected to be small in relation to S. However, the amount of δ_mean_ might be still significant, if the target value was revealed by a more precise reference method. In general, the ratio ν=δ_mean_/S is expected to be clearly non-zero but distinctly smaller than the ratio φ=δ_max_/s_max_

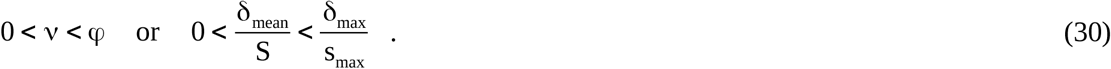

The ratios φ of almost all analytes in clinical chemistry have been revealed and compared in [2,9]. The range (0.6,2.1) covers almost all φ values, where most of the φ values are near 1.0. It is further assumed that all modern and well-established analytic techniques show ratios below 1.5. According to Eq. (30) an adequate value for ν is approx. 0.6 with an assumed maximum uncertainty of about ±0.5. Nevertheless, ν-values of up to 1.5 are considered in this article. Further, the decision to apply a one- or two-sided 95% CI for the parameter z_0.95_ also depends on ν. If the amount of δ_mean_ is almost as high as S (or even higher), a one-sided CI is more suitable than a two-sided CI. Here, a smooth approximate transition between two-sided (ν=0, z=1.96) and one-sided (ν=1, z=1.645) statistics is applied by the sigmoidal function

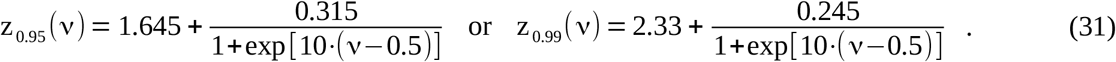

Because Eq. (29) has only two remaining variables, a 3D-plot is provided in Fig. 2 covering the range ν=0-1.5 and n=2-30. Keeping in mind that L_rel_^RMSTD^ goes to 1.0 at very high n, the dependency on ν in the most relevant range 0.1<ν<1.1 to the entire shape of L_rel_(n) is relatively small. The combination of ν=0.6 and z=1.7 will be used as a feasible representative choice for almost all analytes in clinical chemistry.

**Figure 2:**
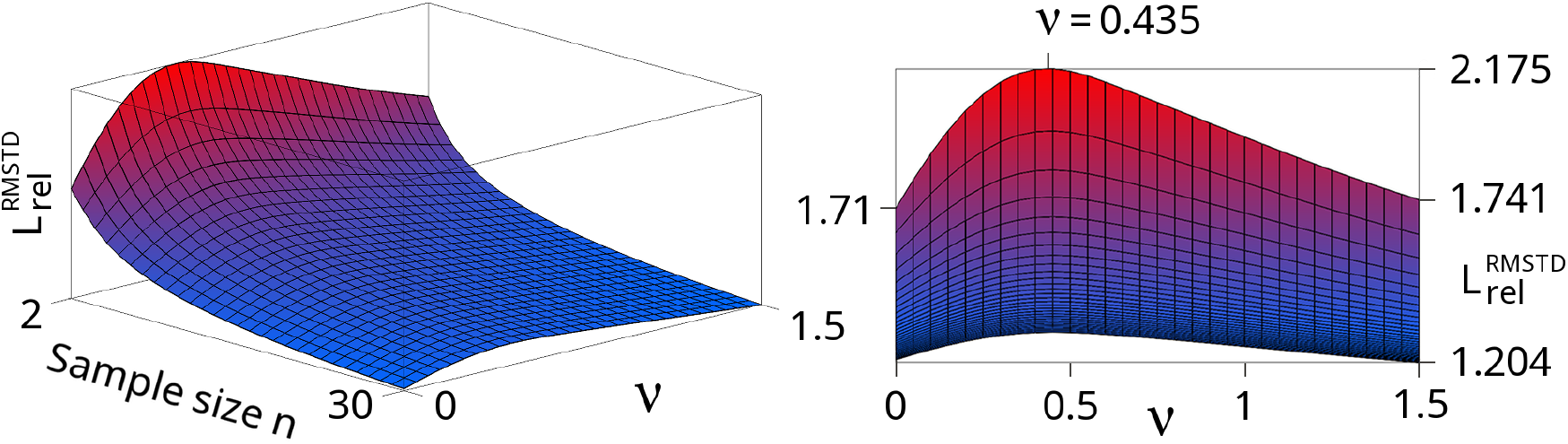
A 3D-plot of the relative upper CI-95% limit of the CI_Δ_^up^ approach is drawn according to Eq. (29). The function is not defined at n=1; thus, the given range of n is 2-30. The considered range of the bias-dispersion-ratio ν is 0-1.5, where the z-value has been adapted with regard to ν (see Eq. (31)). The frontal view at the right-hand side additionally marks key quantities of the L_rel_^RMSTD^ function. The minimum value of 1.204 of the given section is reached at n=30; ν=1.5. The initial amplitudes (at fixed n=2) show a roughly parabolic shape from 1.71 (ν=0) via the maximum value 2.175 at ν=0.435 back to 1.741 (ν=1.5).

At a broad CI of 99%, which would result in a very tolerant IQC limit definition, it is strongly recommended to migrate the derivation of CI_Δ_^up^ from maximum error propagation to the Gaussian error propagation (GEP). In this case, Eq. (29) changes to

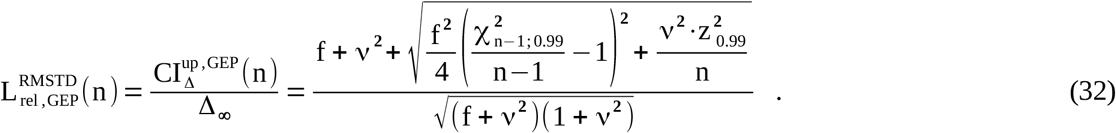

### 3.2 Comparison of both mathematical theories

The more fundamental MDCI theory is comparable to the special case of the CI_Δ_^up^ approach without consideration of an overall bias (ν=0). Thus, both theories are compared in Figs. 3, 4 and 5 utilizing the relative functions given in Eq. (28) and Eq. (29), respectively. Both functions converge towards one at infinity. The intrinsic uncertainty of the CI_Δ_^up^ approach with regard to the concept of the degrees of freedom is indicated by a black/gray corridor. The upper black boundary utilizes the given Eq. (29), whereas the lower black boundary represents the substitution of the term χ _n_^2^/(n-1) by χ_n_ ^2^/n in Eq. (29), which has been discussed in Chapter 2.2. Moreover, the CI_Δ_^up^ approach becomes additionally uncertain at very small sample sizes n<6, mainly due to the approximate correction of the degrees of freedom by the prefactor f. Attempts have been made to introduce a more complex correction of the degrees of freedom [10]. However, the theory behind a fine-tuned adaptation of the prefactor f is beyond the scope of this article.

**Figure 3:**
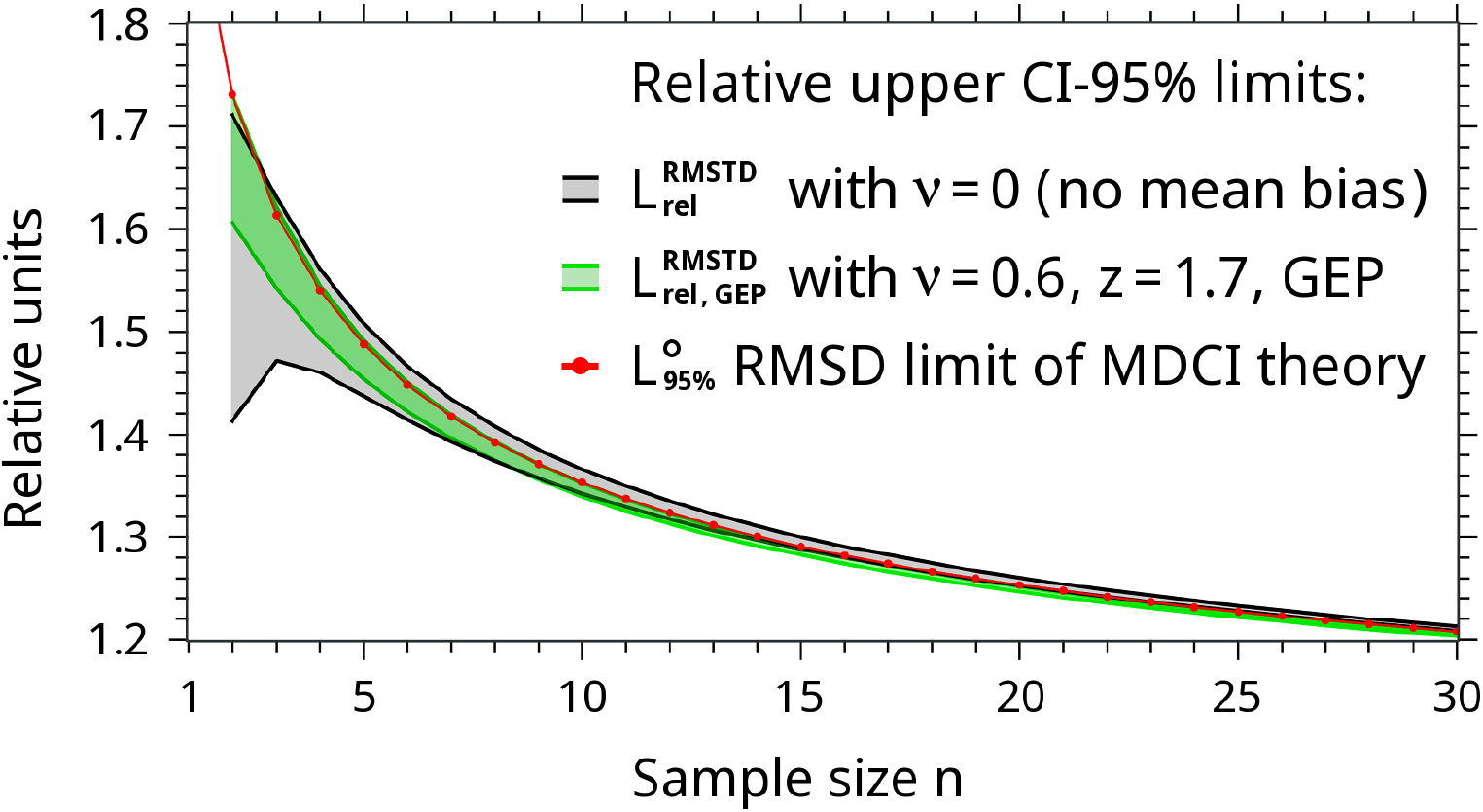
The graph shows the relative increase of the statistical uncertainty of RMSD values with regard to the sample size n at a CI of 95%. The gray corridor and the flanking black graphs are obtained from the CI_Δ_^up^ approach using Eq. (29) with ν=0 (no bias). The red graph represents the MDCI function (Eq. (28)). The narrower green corridor is taken for comparison. It shows the L_rel,GEP_^RMSTD^ corridor obtained by Eq. (32) with ν=0.6 and z=1.7, applying a Gaussian error propagation at a CI of 95%. (For ν=0, as depicted by the gray corridor, the type of error propagation is irrelevant and Eq. (29) = Eq. (32).) All graphs converge to 1 at n→∞. A value for n=1 is not defined by the CI_Δ_^up^ approach.

**Figure 4:**
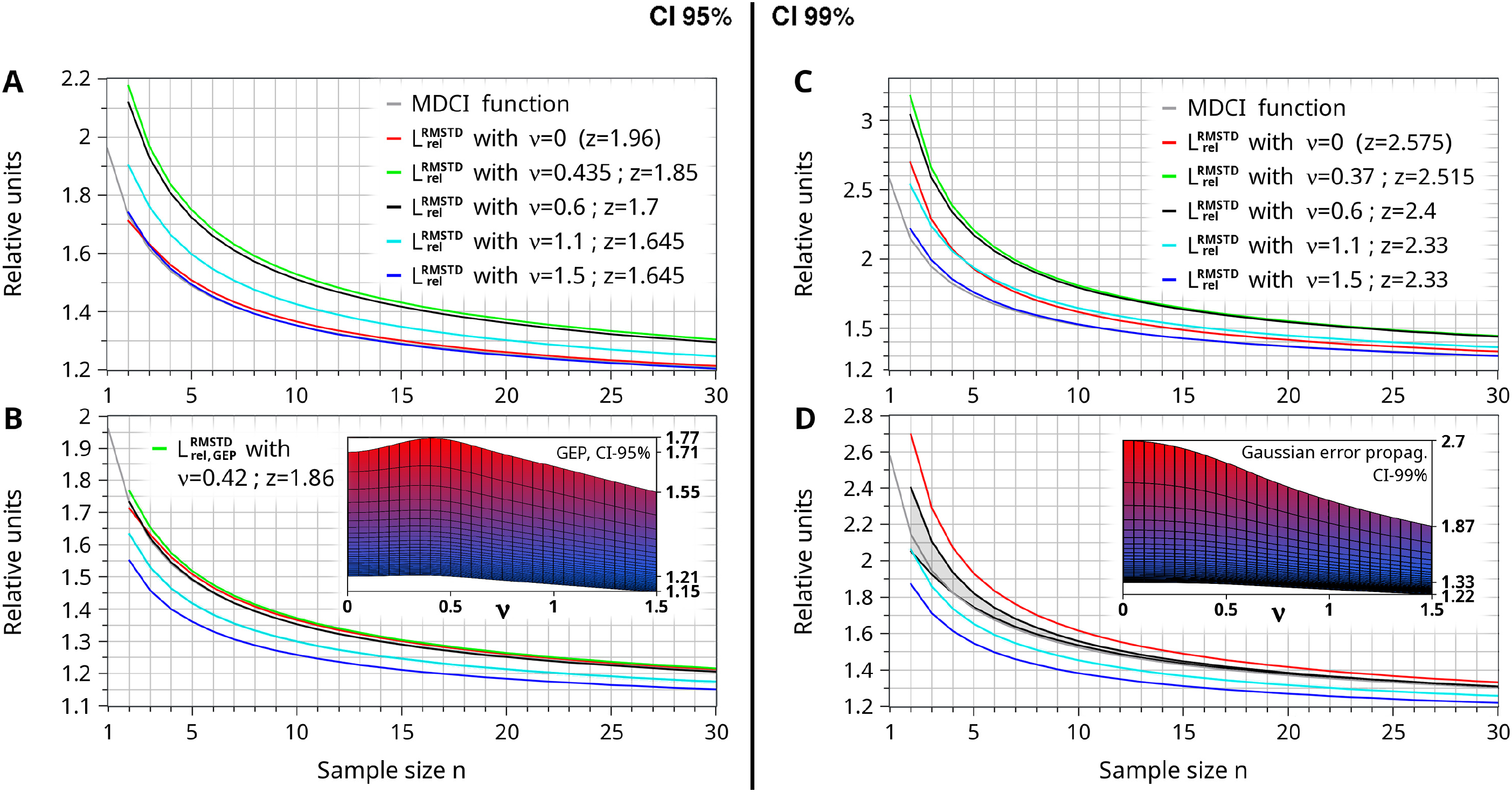
The graphs show the MDCI function (L°, Eq. (28)) and selected curves of the CI_Δ_^up^ approach (L_rel_^RMSTD^, Eq. (29) or L_rel,GEP_^RMSTD^, Eq. (32)) with different values of ν (and dedicated z) within the most relevant range 0≤ν≤1.5. The given functions of the CI_Δ_^up^ approach are usually only the upper boundaries of the shape corridors (as directly obtained by the Eq. (29) / (32)). In (D) the complete L_rel,GEP_^RMSTD^ corridor is exceptionally given for ν=0.6 (black lines). Graphs (A) and (B) are revealed considering a CI of 95%, where (C) and (D) base on a CI of 99%. In vertical direction the graphs differ by the type of the error propagation, which is the maximum error propagation L_rel_^RMSTD^ in (A) and (C) and the Gaussian error propagation L_rel,GEP_^RMSTD^ in (B) and (D), respectively. The color-codes indicate the same values of ν in all four graphs except the green lines, which mark the individual local maximum (if one exists) for ν>0. Frontal views of the entire 3D plots of L_rel,GEP_^RMSTD^ are included as picture-in-picture, where the hidden scale is n=2-30 (see Fig. 2). The frontal view assigned to (A) is given in Fig. 2.

**Figure 5:**
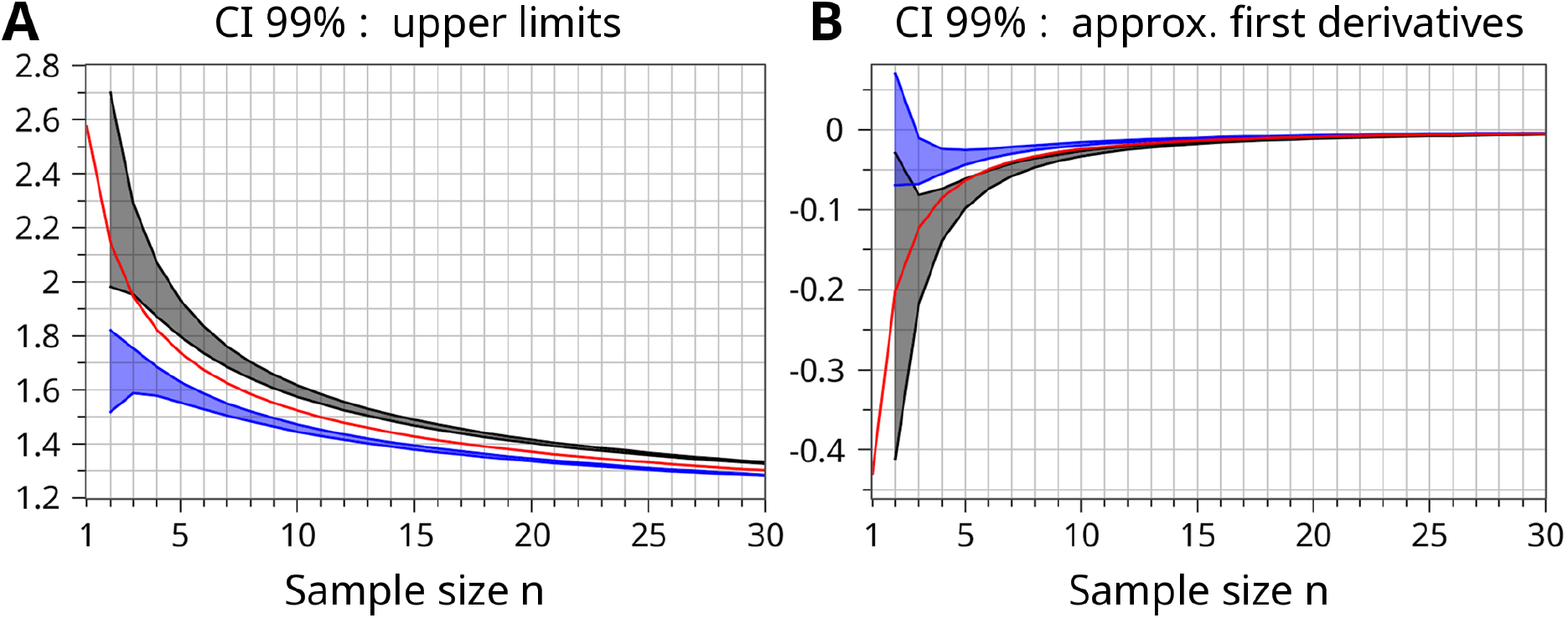
Comparison of the curves of the upper limits of the CI-99%. (A) The amplitudes of the MDCI-99% function (red), the shape corridor of L_rel_^RMSTD^ (ν=0, CI-99%) with variance-based error expansion (black/gray), and the corridor of L_rel_^RMSTD^ (ν=0, CI-99%) with standard-deviation-based error expansion (blue) are compared. (B) The approximate first derivatives of all drawn curves in (A) are compared in addition. The slopes L_rel_^RMSTD^(n+1)-L_rel_^RMSTD^(n) or L°(n+1)-L°(n) have been taken as a simple approximation of the first derivative.

At a CI of 95%, the MDCI function is entirely covered by the shape corridor of L_rel_^RMSTD^ with ν=0 as shown in Fig. 3. It can be further concluded that the upper black curve (representing the unmodified Eq. (29)) is in best agreement with the MDCI function under special consideration of the range 1<n<15. Any in-principle thinkable modification of Eq. (29) (e.g., a more complex prefactor f, a derivation with regard to the standard deviation instead of the variance, a substitution of z by the analog quantile t_n-1_ of the Student distribution) result in significantly higher differences of the entire L_rel_^RMSTD^ corridor to the MDCI function. The remaining discrepancy at very small n can most probably be dedicated to the approximate adaptation of the degrees of freedom. Thus, the CI_Δ_^up^ curvature tends to be slightly less steep at n<6. It is assumed that this conclusion can be generalized to all CI_Δ_^up^ functions (with ν>0). The question, why the relative CI_Δ_^up^ function at ν=0 is not fully identical to the MDCI function, is addressed in a short appendix of this article.

The green corridor has been added to Fig. 3 for comparison. It marks the shape corridor of the CI_Δ_^up^ approach (L_rel,GEP_^RMSTD^) utilizing the parameter set ν=0.6 and z_0.95_=1.7 (which is considered as representative for analytes in clinical chemistry) in combination with the Gaussian error propagation according to Eq. (32) at a CI of 95%. Surprisingly, the agreement between the upper green boundary of L_rel_^RMSTD^ and the red MDCI-95% function is nearly perfect. This agreement is also visualized in Fig. 4B as black and gray lines. Further, the green corridor in Fig. 3 is equivalent to the black corridor in Fig. 4D except the different CI. Applying the maximum error propagation, the MDCI-95% function is still within the corridor of L_rel_^RMSTD^ with ν=0.6, z_0.95_=1.7 as shown in [2].

The situation at a CI of 99% is less clear, because the CI_Δ_^up^ functions increase more rapidly than the MDCI function, going from CI-95% to CI-99%. A comparison is shown in Fig. 4 and discussed below. Further information are provided in the appendix and Fig. 5. Nevertheless, the L_rel,GEP_^RMSTD^ corridor according to Eq. (32) with ν=0.6 and z_0.99_=2.4 comes again in agreement with the shape of the corresponding MDCI-99% function. The comparison is illustrated in Fig. 4D and [2].

### 3.3 Conclusions with regard to small sample sizes

The study quantitatively revealed that a significant statistical uncertainty in the determination of RMSTD or RMSD metrics must be considered if the sample size (n) is below about 50. This uncertainty (or error) can reach additional 100% (or even more) of the true RMS(T)D value. Two mathematical approaches were derived to quantify this uncertainty. The CI_Δ_^up^ approach refers to the entire RMSTD (inclusive bias); however, it lacks precision below about n<6. The second approach (MDCI) represents a more fundamental theory, but it neglects a mean bias with respect to a target value (just RMSD). Both final functions - the relative CI_Δ_^up^ function L_rel_^RMSTD^ (Eq. (29)) and the MDCI function L° (Eq. (28)) - define n-dependent prefactors to the true values of RMSTD or RMSD. Despite differences in the final equations, the L_rel_^RMSTD^ function without bias (ν=0) is very similar to the MDCI function if a CI of 95% is considered, as shown in Figs. 3 and 4.

At very broad CI ranges (CI=99%) the comparison of both approaches is less clear as discussed in Chapter 3.2. and the appendix. As shown in Fig. 4, the MDCI functions generally stay in good agreement with the L_rel,GEP_^RMSTD^ (ν=0.6) corridors at both CI-99% and CI-95%, if the CI_Δ_^up^ approach is switched from maximum to Gaussian error propagation. Nevertheless, a CI significantly above 95% is not recommended. For IQC a combination of a rather narrow CI of about 95% in addition to a small constant offset to all evaluated maximum permissible limits is favorable. This small general bonus allows a slightly extended basic tolerance for limited baseline variation. The offset might be in the order of the difference in maximum amplitudes (at n=2) between the CI-99% and CI-95% functions.

The quantification of the statistical uncertainty allows to optimally adapt general IQC limits for RMSTD results with regard to the sample size of a particular SMC chart or to the size of a retrospective monitoring window of SMC data. More precisely, both revealed functions L° and L_rel_^RMSTD^ represent an adaptation function a(n) to consider limited sample sizes - leading to the n-dependent IQC limit L(n)=a(n)·L(∞). In [2] a simple fit function for a(n) in the range n<30 is suggested. The presented findings led to the development of a novel powerful IQC method [2] that efficiently monitors a very limited available number of recent SMC values.

Established quantitative diagnostic methods in clinical chemistry provide mean-bias-to-overall-deviation ratios in the range 0<ν<1.1. This conclusion is extensively discussed in [2]. The most populated region is expected to be ν≈0.6±0.3. Thus, the black curves (ν=0.6) in Fig. 4 indicate the representative functions for all analytes in clinical chemistry. The red (ν=0) and cyan (ν=1.1) curves mark the approximate boundaries of realistic ratios in clinical chemistry.

### 3.4 Risks of a short evaluation period in combination with Westgard Sigma Rules

The “Westgard Sigma Rules” is currently a very prominent IQC approach in clinical chemistry. It is a serial multirule concept starting with the 3S entry rule. This rule defines a first in-control range of μ_e_±3·S_e_ also known as the Shewhart rule. Both parameters (mean value μ_e_ and standard deviation S_e_) must be determined during a pre-analytical evaluation period, if the control sample is unlabeled. First, we assume normal-distributed SMC values during the analytic process and a negligible bias (the means of dedicated SMC charts are almost similar to μ_e_). The 3S range thus covers 99.9% of the expected data, and the rule causes only about 0.2% false alerts. This sounds comfortable. However, a problem arises if the parameter S_e_ was insufficiently determined due to small amounts of evaluation data. Several laboratories only use 20 data points during evaluation. Let us define σ as the unknown real in-control standard deviation. The limits ±3·σ would provide the above mentioned false-alert rate of only 0.2%. Considering 20 data points, the risk is high that the real standard deviation has been significantly underestimated. In an unfavorable but still realistic scenario, the obtained S_e_ is near or below the lower limit of the two-sided 95% CI around σ. This happens on average in 1 of 40 evaluation processes. According to Eqs. (12) and (13), the CI^low^ is given by

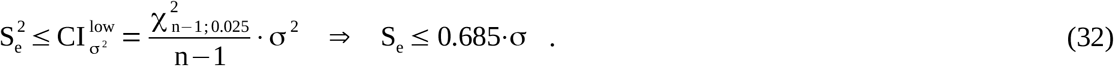

Applying this unwittingly underestimated standard deviation, Westgard’s 3S entry rule actually becomes a 2S rule

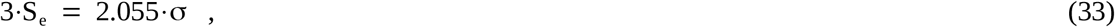

and the rate of false alerts increases to 4%. Thus, on average, one false alert every 25 IQC tests must be expected in this situation. Further, the expectation value μ_e_ has also been self-determined with a distinct uncertainty: 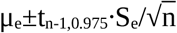 (t_n-1,0.975_: 95%-quantile of the Student distribution). At n=20 the uncertainty of the revealed mean would be μ_e_±0.47·S_e_. The resulting distortion of the real mean further increases the problem of false alerts. It is important to note that all mentioned uncertainties are purely statistical effects. They can not be circumvented by consideration of very different operation conditions during the evaluation period. In fact, the evaluated standard deviation S_e_ would be even more underestimated (compared to the true σ), if the considered evaluation data not entirely reflect the full spectrum of potential operation conditions.

A pragmatic solution to ensure a sufficient evaluation of μ_e_ and S_e_ would be a share of peer-group data combined with an ANOVA analysis or the approach described in Chapter 2.1. A solely self-made parameter evaluation should ideally base on at least 40 values, which would reduce the final rate of false alerts of the 3S rule to below 1.9%. A more tolerant entry rule (e.g., ±3.5S or ±4S limits) of the Westgard Sigma Rules might also be part of a practicable solution, which would be particularly suitable for techniques with a high Sigma Metric.

## Data Availability

All data sources are mentioned in the reference list.

## Abbreviations

ANOVA: analysis of variance
CI: confidence interval
CI_Δ_^up^: upper limit of the confidence interval of the RMSTD metric
Δ: the “true” RMSTD value determined using sufficient amounts of data
GEP: Gaussian error propagation
GPD: Gaussian probability density
IQC: internal quality control
MDCI: multidimensional confidence interval
RA: retrospective (statistical) analysis of SMC data
RMSD: root mean square deviation (aka empirical standard deviation)
RMSTD: root mean square total (or target) deviation with respect to a known target value (aka entire analytical measuring uncertainty)
SMART: Statistical Monitoring by Adaptive RMSTD Tests
SMC: single measurement of a control sample

## Appendix

How does the CI_Δ_^up^ approach at ν=0 (no bias) must be modified to become identically equal to the MDCI function? First, all modifications with regard to a correction of the degrees of freedom are rejected (esp. the prefactor f is neglected). Finally, the error propagation leading to CI_Δ_^up^ has to utilize the standard deviation S instead of the variation S^2^. Please note that a derivation considering the variation has been preferred because (in contrast to the square root of S^2^) the estimator S^2^=1/(n-1)·∑(y_i_-<y>)^2^ is unbiased.

Utilizing S, the derivative and uncertainty (according to Eq. (8) and (14)) change to

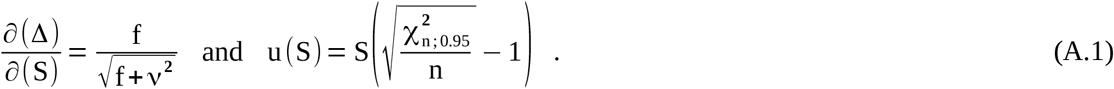

Thus, Eq. (16) becomes to

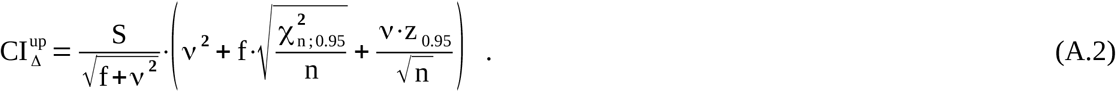

The resulting relative function L_rel_^RMSTD^ finally ends up in the MDCI function if f=1 and ν=0.

Based on a CI of 95%, the MDCI function lies within the shape corridor of L_rel_^RMSTD^ (always at ν=0) obtained by the preferred derivation via the error of the variance (see Eq. (29) and Fig. 3). The flanking functions of any L_rel_^RMSTD^ corridor differ by the term χ _(n-1)_^2^/(n-1) (upper curve) or χ _n_^2^/n instead (lower curve), as discussed in Chapter 2.2. The corresponding corridor of the alternate derivation of CI_Δ_^up^ via the error of the standard deviation (as given in Eq. (A.2) above) lies significantly below (not shown for CI-95%).

Based on a CI of 99%, the increases of both L_Δ_^RMSTD^ corridors towards higher amplitudes are stronger than that of the MDCI function; thus, the MDCI function now appears between both corridors. This constellation is drawn in Fig. 5 for the sake of completeness. Please note that the CI_Δ_^up^ approach is based on just a first-order Taylor series expansion. It is assumed that the approach becomes less accurate at higher amplitudes (i.e., at higher CI levels). The corridors in Fig. 5B further demonstrate that almost all kinds of CI_Δ_^up^ derivations clearly fail at n≤4, most probably due to the simplified handling of the degrees of freedom.

## Research Funding

This research did not receive any specific grant from funding agencies in the public, commercial, or not-for-profit sectors.

---

**Table S1:** Relevant metrics Γ(n/2), z(n), and L°(n) for all n≤40 and at the two-sided CI levels 95%, 97,5%, and 99%.

(n: dimension or sample size. Values $z(n)$ are equivalent to the square root of the appropriate chi-square value.)
| n | Gamma(n/2) | z(95%) | RMSD°(95%) | z(97.5%) | RMSD°(97.5%) | z(99%) | RMSD°(99%) |
| --- | --- | --- | --- | --- | --- | --- | --- |
| 1 | 0.177245E+01 | 1.959939 | 1.959939 | 2.241378 | 2.241378 | 2.575804 | 2.575804 |
| 2 | 0.100000E+01 | 2.447722 | 1.730801 | 2.716178 | 1.920628 | 3.034829 | 2.145948 |
| 3 | 0.886227E+00 | 2.795458 | 1.613959 | 3.057491 | 1.765243 | 3.368189 | 1.944625 |
| 4 | 0.100000E+01 | 3.080191 | 1.540095 | 3.338131 | 1.669066 | 3.643696 | 1.821848 |
| 5 | 0.132934E+01 | 3.327211 | 1.487974 | 3.582223 | 1.602019 | 3.884080 | 1.737013 |
| 6 | 0.200000E+01 | 3.548438 | 1.448644 | 3.801208 | 1.551837 | 4.100206 | 1.673902 |
| 7 | 0.332335E+01 | 3.750594 | 1.417591 | 4.001570 | 1.512451 | 4.298266 | 1.624592 |
| 8 | 0.600000E+01 | 3.937908 | 1.392261 | 4.187402 | 1.480470 | 4.482188 | 1.584693 |
| 9 | 0.116317E+02 | 4.113243 | 1.371081 | 4.361485 | 1.453828 | 4.654649 | 1.551550 |
| 10 | 0.240000E+02 | 4.278647 | 1.353027 | 4.525809 | 1.431187 | 4.817573 | 1.523450 |
| 11 | 0.523428E+02 | 4.435642 | 1.337396 | 4.681860 | 1.411634 | 4.972396 | 1.499234 |
| 12 | 0.120000E+03 | 4.585394 | 1.323689 | 4.830779 | 1.394526 | 5.120226 | 1.478082 |
| 13 | 0.287885E+03 | 4.728826 | 1.311540 | 4.973465 | 1.379391 | 5.261938 | 1.459399 |
| 14 | 0.720000E+03 | 4.866677 | 1.300674 | 5.110645 | 1.365877 | 5.398237 | 1.442740 |
| 15 | 0.187125E+04 | 4.999554 | 1.290879 | 5.242912 | 1.353714 | 5.529705 | 1.427764 |
| 16 | 0.504000E+04 | 5.127960 | 1.281990 | 5.370762 | 1.342690 | 5.656823 | 1.414206 |
| 17 | 0.140344E+05 | 5.252319 | 1.273874 | 5.494610 | 1.332639 | 5.780000 | 1.401856 |
| 18 | 0.403200E+05 | 5.372991 | 1.266426 | 5.614811 | 1.323424 | 5.899577 | 1.390544 |
| 19 | 0.119292E+06 | 5.490287 | 1.259558 | 5.731670 | 1.314935 | 6.015859 | 1.380133 |
| 20 | 0.362880E+06 | 5.604476 | 1.253199 | 5.845452 | 1.307083 | 6.129105 | 1.370510 |
| 21 | 0.113328E+07 | 5.715768 | 1.247283 | 5.956365 | 1.299785 | 6.239516 | 1.361574 |
| 22 | 0.362880E+07 | 5.824419 | 1.241770 | 6.064660 | 1.292990 | 6.347340 | 1.353257 |
| 23 | 0.118994E+08 | 5.930587 | 1.236613 | 6.170495 | 1.286637 | 6.452732 | 1.345488 |
| 24 | 0.399168E+08 | 6.034437 | 1.231774 | 6.274030 | 1.280681 | 6.555850 | 1.338207 |
| 25 | 0.136843E+09 | 6.136112 | 1.227222 | 6.375408 | 1.275082 | 6.656834 | 1.331367 |
| 26 | 0.479002E+09 | 6.235745 | 1.222930 | 6.474760 | 1.269805 | 6.755813 | 1.324924 |
| 27 | 0.171054E+10 | 6.333454 | 1.218874 | 6.572203 | 1.264821 | 6.852901 | 1.318841 |
| 28 | 0.622702E+10 | 6.429346 | 1.215032 | 6.667843 | 1.260104 | 6.948204 | 1.313087 |
| 29 | 0.230923E+11 | 6.523520 | 1.211387 | 6.761776 | 1.255630 | 7.041816 | 1.307632 |
| 30 | 0.871783E+11 | 6.616064 | 1.207923 | 6.854090 | 1.251380 | 7.133825 | 1.302452 |
| 31 | 0.334839E+12 | 6.707061 | 1.204624 | 6.944868 | 1.247335 | 7.224311 | 1.297524 |
| 32 | 0.130767E+13 | 6.796586 | 1.201478 | 7.034184 | 1.243480 | 7.313348 | 1.292829 |
| 33 | 0.519000E+13 | 6.884707 | 1.198474 | 7.122104 | 1.239799 | 7.400999 | 1.288349 |
| 34 | 0.209228E+14 | 6.971490 | 1.195601 | 7.208695 | 1.236281 | 7.487334 | 1.284067 |
| 35 | 0.856350E+14 | 7.056992 | 1.192849 | 7.294012 | 1.232913 | 7.572404 | 1.279970 |
| 36 | 0.355687E+15 | 7.141270 | 1.190212 | 7.378113 | 1.229685 | 7.656268 | 1.276045 |
| 37 | 0.149861E+16 | 7.224375 | 1.187680 | 7.461047 | 1.226589 | 7.738974 | 1.272279 |
| 38 | 0.640237E+16 | 7.306354 | 1.185247 | 7.542862 | 1.223614 | 7.820569 | 1.268664 |
| 39 | 0.277243E+17 | 7.387252 | 1.182907 | 7.623601 | 1.220753 | 7.901096 | 1.265188 |
| 40 | 0.121645E+18 | 7.467110 | 1.180654 | 7.703307 | 1.218000 | 7.980599 | 1.261843 |

